# Waning of SARS-CoV-2 antibodies targeting the Spike protein in individuals post second dose of ChAdOx1 and BNT162b2 COVID-19 vaccines and risk of breakthrough infections: analysis of the Virus Watch community cohort

**DOI:** 10.1101/2021.11.05.21265968

**Authors:** Robert W Aldridge, Alexei Yavlinsky, Vincent Nguyen, Max T Eyre, Madhumita Shrotri, Annalan M D Navaratnam, Sarah Beale, Isobel Braithwaite, Thomas Byrne, Jana Kovar, Ellen Fragaszy, Wing Lam Erica Fong, Cyril Geismar, Parth Patel, Alison Rodger, Anne M Johnson, Andrew Hayward, on behalf of the Virus Watch study

**Affiliations:** Centre for Public Health Data Science, Institute of Health Informatics, University College London, UK; Institute of Epidemiology and Health Care, University College London, London, UK; Centre of Health Informatics, Computing and Statistics, Lancaster Medical School, Lancaster University, Lancaster, LA1 4YW, UK; Liverpool School of Tropical Medicine, Liverpool, UK; Department of Infectious Disease Epidemiology, London School of Hygiene and Tropical Medicine, Keppel Street, London, UK; Institute for Global Health, University College London, London, UK

## Abstract

**Background:** SARS-CoV-2 vaccines stimulate production of antibodies targeting the spike protein (anti-S). The level of antibodies following vaccination and trajectories of waning may differ between vaccines influencing the level of protection, how soon protection is reduced and, consequently the optimum timing of booster doses.

**Methods:** We measured SARS-CoV-2 anti-S titre in the context of seronegativity for SARS-CoV-2 anti-Nucleocapsid (anti-N), in samples collected between 1st July and 24th October 2021 in a subset of adults in the Virus Watch community cohort. We compared anti-S levels after BNT162b2 (BioNTech/Pfizer) or ChAdOx1 (AstraZeneca/Oxford) vaccination using time since second dose of vaccination, age, sex and clinical vulnerability to investigate antibody waning. To investigate the use of anti-S levels as a correlate of protection against SARS-CoV-2 infection, we undertook a survival analysis (Kaplan-Meier and Cox) with individuals entering 21 days after their second dose of vaccine, or first antibody test after 1st July (whichever was latest) and exiting with the outcome of SARS-Cov-2 infection or at the end of follow up 24th October 2021. We also undertook a negative test design case-control analysis of infections occurring after the second vaccine dose (breakthrough infections) to determine whether the type of vaccine affected the risk of becoming infected.

**Results:** 24049 samples from 8858 individuals (5549 who received a second dose of ChAdOx1 and 3205 BNT162b2) who remained anti-N negative were included in the analysis of anti-S waning over time. Three weeks after the second dose of vaccine BNT162b2 mean anti-S levels were 9039 (95%CI: 7946-10905) U/ml and ChadOx1 were 1025 (95%CI: 917-1146) U/ml. For both vaccines, waning anti-S levels followed a log linear decline from three weeks after the second dose of vaccination. At 20 weeks after the second dose of vaccine, the mean anti-S levels were 1521 (95%CI: 1432-1616) U/ml for BNT162b2 and 342 (95%CI: 322-365) U/ml for ChadOx1. We identified 197 breakthrough infections and found a reduced risk of infection post second dose of vaccine for individuals with anti-S levels greater than or equal to 500 U/ml compared to those with levels under 500 U/ml (HR 0.62; 95%CIs:0.44-0.87; p=0.007). Time to reach an anti-S threshold of 500 U/ml was estimated at 96 days for ChAdOx1 and 257 days for BNT162b2. We found an increased risk of a breakthrough infection for those who received the ChAdOx1 compared to those who received BNT162b2 (OR: 1.43, 95% CIs:1.18-1.73, p<0.001).

**Discussion:** Anti-S levels are substantially higher following the second dose of BNT162b2 compared to ChAdOx1. There is a log linear waning in levels for both vaccines following the second dose. Anti-S levels are an important correlate of protection as demonstrated by those with anti-S levels < 500U/ml following vaccination being at significantly greater risk of subsequent infection. Since anti-S levels are substantially lower in ChAdOx1 than in BNT162b2 and both decline at similar rates we would expect waning immunity to occur earlier in ChAdOx1 compared to BNT162b2. Our results showing an increased risk of breakthrough infections for those who were vaccinated with ChAdOx1 compared to BNT162b2 are in line with this hypothesis. Consistent with our data, national analyses of vaccine effectiveness also suggest that waning of immunity for infection and, to a lesser extent for severe disease, is seen earlier in ChAdOx1 than in BNT162b2. Our data demonstrate the importance of booster doses to maintain protection in the elderly and clinically vulnerable and suggest that these should be prioritised to those who received ChAdOx1 as their primary course.

## Introduction

Vaccines based on the spike glycoprotein of SARS-CoV-2 are estimated to have prevented the deaths of hundreds of thousands of people globally.^7–9^ On 1 November 2021, 50,025,020 people in the UK had received their first dose of COVID-19 vaccine, 45,731,565 had received a second dose and 8,356,172 people had received a booster or third dose.^10^ The majority of people aged over 18 in the UK were vaccinated with either BNT162b2 (BioNTech/Pfizer) or ChAdOx1 nCoV-19 (AstraZeneca/Oxford).^11^ We have previously reported^12^ an analysis of waning SARS-CoV-2 antibodies targeting the spike protein (anti-S) after the second dose of BNT162b2 or ChAdOx1 in 605 adults with samples collected on June 14–15 2021. These data suggested higher peak levels and faster waning of anti-S levels after a second dose of BNT162b2 compared to ChAdOx1 in infection-naive individuals over a 3–10-week period.

Recent analysis by the UK Health Security Agency (UKHSA)^1^ estimates that 20 weeks following the second dose, vaccine effectiveness for ChAdOx1 against infection was 47.3% (95% CIs:45.0-49.6) compared to 69.7% (95% CIs: 68.7-70.5) for BNT162b2. For hospitalisations due to COVID-19, the corresponding vaccine effectiveness was 77.0% (70.3-82.3) for ChadOx1 and 92.7% (90.3-94.6) for BNT162b2. For deaths, vaccine effectiveness at 20 weeks was 78.7 (95% CI 52.7 to 90.4) for ChadOx1and 90.4 (95% CI 85.1 to 93.8) for BNT162b2. Overall, these data suggest faster waning of protection against infection and severe disease for ChAdOx1 compared to BNT162b2.

Transmission of SARS-CoV-2 remains high in the UK with 33,865 people tested positive, 1,002 patients admitted to hospital and 293 deaths within 28 days of positive test reported on 2 November 2021.^10^ Between 27th September 2021 and 24th October 2021, 96% of deaths occurring within 28 days of a positive SARS-CoV-2 test were in those aged 50 and over and over half were in those aged 80 and above. Although rates of death are substantially lower in vaccinated groups, 79% of these deaths were in those who had been double vaccinated at least 14 days prior to the positive SARS-CoV-2 test (and 87% in those aged 80 years and above).^13^

Given the high levels of vaccination in the UK and consequently the high level of cases and deaths occurring in double vaccinated individuals, it is important to understand trajectories of anti-S waning in individuals post second dose of COVID-19 vaccine and whether they can be used as a correlate of protection for SARS-CoV-2 infection, to enable serosurveillance data to inform vaccine policies around the globe. In this analysis we aim to build upon our prior evidence of anti-S waning and directly compare the effectiveness of BNT162b2 and ChAdOx1 in preventing SARS-CoV-2 infection in the Virus Watch cohort. We examine three research questions. First, how does waning vary by vaccine type, demographic and clinical characteristics. Second, is there evidence that the anti-S level is a meaningful correlate of protection against SARS-Cov-2 infection and what is the predicted time to reach a specified anti-S level potentially representing a threshold of protection for each of the 2 commonly used vaccines in the UK. Third, how does the type of vaccine affect the chances of developing COVID-19 after receiving two vaccine doses.

## Methods

The Virus Watch study is a household community cohort of acute respiratory infections in England & Wales that started recruitment in June 2020.^6^ A detailed description of recruitment methods has been described previously^6^, but in summary, to recruit our sample we used a range of methods. We used the Royal Mail Post Office Address File to generate a random list of residential address lists that were sent recruitment postcards, we placed social media adverts on Facebook and Twitter and sent SMS messages and letters to participants from their General Practitioners. Participants were followed-up weekly by email with a link to an illness survey which asked about the presence or absence of symptoms that could indicate COVID-19 disease including respiratory, gastrointestinal and general infection symptoms, in addition to cold-like symptoms such as headache, sore throat and runny nose. The weekly survey was also used to capture SARS-CoV-2 test results received from outside the study (eg. via the UK Test-Trace-Isolate system).

### SARS-CoV-2 Antibody testing

Nested within the larger Virus Watch study is a sub-cohort of 10,330 adults (aged over 18) participating in antibody testing who completed at-home capillary blood sampling kits sent via post on a monthly basis. We measured antibody titres targeting the spike (S) protein (anti-S) in the context of seronegativity for SARS-CoV-2 anti-Nucleocapsid (anti-N) which is associated with natural infection. Sera were tested using Elecsys anti-S and anti-N electro-chemiluminescent immunoassays (Roche Diagnostics, Basel, Switzerland).^14^ The anti-S assay targets total antibodies to the S1 subunit of the spike protein (range 0·4–25 000 units per mL [U/mL]), whereas the anti-N assay targets total antibodies to the full length nucleocapsid protein, which we took as a proxy for previous SARS-CoV-2 infection (specificity 99·8% [99·3–100]).^15^ Individuals were included in this analysis if they underwent antibody testing (anti-N and anti-S) and had a valid result between 1st July 2021 and 24th October 2021. Antibody results were excluded after individuals reported a third booster COVID-19 vaccination.

### SARS-CoV-2 Antigen testing

We examined SARS-CoV-2 positive tests confirmed using polymerase chain reaction (PCR) or rapid lateral flow antigen tests (LFD). These positive tests were identified either by participant self-report (1st July 2020 to 24th October 2021) or from linkage of patient demographic characteristics (name, date of birth, address, NHS number) to the national Second Generation Surveillance System for SARS-CoV-2 from 1st July 2020 to 11th August 2021.

### Outcomes

We considered two primary outcomes. First, SARS-CoV-2 anti-S titre in the context of seronegativity for SARS-CoV-2 anti-N. Second, SARS-CoV-2 positive tests confirmed using PCR or rapid lateral flow antigen tests. We defined breakthrough SARS-CoV-2 infection as a positive test (PCR or LFD) after being fully vaccinated at least 14 days following the second dose of BNT162b2 or ChAdOx1. We only included individuals with anti-S levels measured at least 14 days prior to breakthrough infection to ensure anti-S levels used upon cohort entry were not inflated by early asymptomatic breakthrough infection. For this current analysis, we did not examine the presence or absence of symptoms in the context of a positive SARS-CoV-2 test.

### Covariates

We included self-reported vaccination status, and vaccine type, collected weekly from 11 January 2021 onwards. Age, sex, ethnicity and geographical region were derived from participants’ responses to demographic questions at study baseline. We categorised people as clinically vulnerable (CV) or clinically extremely vulnerable (CEV) using our previously reported methods.^12^ People were considered clinically extremely vulnerable using criteria set out by Public Health England and the Department of Health and Social Care as part of the guidance for shielding^16^, which were adapted in line with clinical variables collected through the Virus Watch baseline survey and a monthly survey. People were categorised as clinically vulnerable (CV) using criteria set out by the Joint Committee on Vaccination and Immunisation on 30 December 2020^17^, but excluding those who met the superseding clinically extremely vulnerable criteria (see supplementary appendix for details). Individuals were classified as not clinically extremely vulnerable, or clinically vulnerable if they did not meet these clinical criteria. We also included individuals with missing data on clinical characteristics as not clinically extremely vulnerable, or clinically vulnerable after review of antibody levels in this group.

### Statistical Analyses

We undertook three separate analyses. First, to investigate antibody waning we compared anti-S levels for ChAdOx1 and BNT162b2 by time since vaccination, age, sex and clinical vulnerability. For each week since vaccination, we calculated the geometric mean of the anti-S levels and associated 95% confidence intervals. We fit a linear mixed effect model with a random intercept (to account for repeated samples for each participant) to anti-S level data in natural log space 21 days post second vaccination dose (to account for increasing anti-S levels prior to this time point) and predicted the trajectory of waning by vaccine type and ChAdOx1 and BNT162b2. We used the estimated regression coefficient *r* from the linear model to calculate the corresponding half-life value associated with anti-S waning, using the formula:

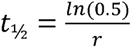

Second, to investigate the use of anti-S levels as a correlate of protection, we undertook a survival analysis using a Kaplan-Meier descriptive analysis in addition to a Cox regression model with individuals entering at risk of SARS-CoV-2 infection 14 days after an antibody test from 1st July and exiting at the first of reported episode of SARS-CoV-2 breakthrough infection or at the end of study follow up on 24th October 2021. Quartiles of anti-S levels were calculated based on samples collected between 1st July 2021 - 30th July 2021. Based on this analysis, we defined an antibody threshold where there was measurable waning in protection and then used log linear projections to predict the timing at which antibody levels would drop to levels associated with decreased protection following the second dose of ChAdOx1 and BNT162b2.

Third, to investigate how the vaccine type affects the chances of developing a breakthrough infection after the second dose, we undertook a negative-test design case-control study in the full Virus Watch cohort (e.g. not just those participants undertaking monthly antibody testing). Cases were defined as double-vaccinated individuals who had not reported prior infection, reporting a PCR or rapid antigen confirmed infection at least two weeks after the second dose of the vaccine and before their booster dose, if applicable. Controls were defined as double-vaccinated individuals who had not reported infection throughout the study period, reporting a negative PCR or rapid antigen test result at least two weeks after the second dose of the vaccine and prior to their booster dose, if applicable. Four controls were matched to each case based on COVID-19 incidence levels in their lower tier local authority at the time of test. The exposure of interest was the type of vaccine (ChAdOx1 or BNT162b2) and the chosen covariates were age, sex, clinical vulnerability and time since receiving the second vaccine dose. Logistic regression was used to calculate the odds ratio for the exposure of interest in the presence of the covariates.

### Ethics

The Virus Watch study has been approved by the Hampstead NHS Health Research Authority Ethics Committee. Ethics approval number - 20/HRA/2320.

## Results

24049 samples from 8858 anti-N negative individuals were included in the analysis of anti-S waning over time (Table 1). 5549 people (63%; 5549/8858) who received a second dose of ChAdOx1 and 3205 who received a second dose BNT162b2 (36%; 3205/8858) were included in our analyses of the trajectories of anti-S waning after the second dose by vaccine type. For both vaccines, waning anti-S levels followed a log linear decline from three weeks after the second dose of vaccination (Figure 1). Three weeks after the second dose of vaccine BNT162b2, mean anti-S levels were 9039 (95%CI: 7946-10905) U/ml and ChAdOx1 were 1025 (95%CI: 917-1146) U/ml. At 20 weeks after the second dose of vaccine, the mean anti-S levels were 1521 (95%CI: 1432-1616) U/ml for BNT162b2 and 342 (95%CI: 322-365) U/ml for ChAdOx1. There was evidence that rates of waning were higher in BNT162b2 (−8.27e-03 [ln(anti-S U/ml)/day], *t*_½_ = 68.81 days) than ChAdOx1 (−10.1e-03 [ln(anti-S U/ml)/day], *t*_½_ = 84.5 days; p<0.001).

**Table 1.** Demographic and clinical characteristics of included individuals and samples.

|  | Individuals | Samples |
| --- | --- | --- |
|  | N (%) | N (%) |
| All | 8,858 (100%) | 24,049 (100%) |
| Age group |  |  |
| 18-64 | 4,574 (52%) | 12,447 (52%) |
| 65+ | 4,284 (48%) | 11,602 (48%) |
| Sex |  |  |
| Male | 3,749 (42%) | 10,163 (42%) |
| Female | 5,098 (58%) | 13,858 (58%) |
| Missing/Other | 11 (0.1%) | 28 (0.1%) |
| Ethnicity |  |  |
| White British or Irish | 8,240 (93%) | 22,517 (94%) |
| White Other | 365 (4.1%) | 912 (3.8%) |
| Mixed | 63 (0.7%) | 157 (0.7%) |
| South Asian | 85 (1.0%) | 194 (0.8%) |
| Other Asian | 48 (0.5%) | 127 (0.5%) |
| Black | 23 (0.3%) | 59 (0.2%) |
| Other/Missing | 34 (0.4%) | 83 (0.3%) |
| Clinical vulnerability |  |  |
| Clinically extremely vulnerable | 1,247 (14%) | 3,366 (14%) |
| Clinically vulnerable | 2,399 (27%) | 6,502 (27%) |
| Not clinically vulnerable | 5,212 (59%) | 14,181 (59%) |
| 2nd dose vaccine type |  |  |
| BNT162b2 | 3,205 (36%) | 8,132 (34%) |
| ChAdOx1 | 5,549 (63%) | 15,659 (65%) |
| mRNA-1273 | 70 (0.8%) | 169 (0.7%) |
| Other/ Missing | 34 (0.4%) | 89 (0.4%) |

See supplementary appendix for further details of clinically vulnerable and clinically extremely vulnerable groups.

**Figure 1.**
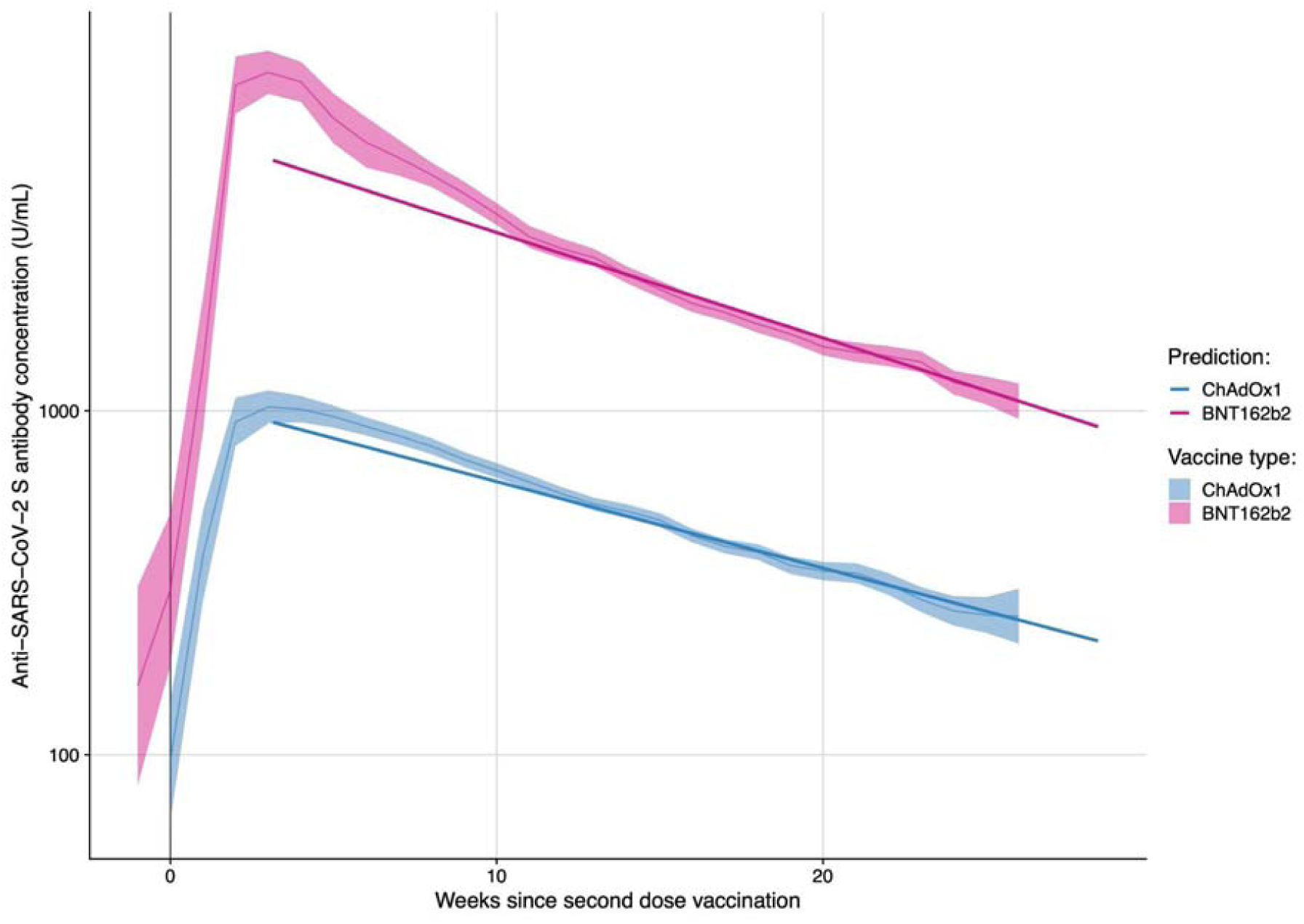
Anti-S levels (U/ml) over time since second dose of vaccination amongst N-seronegative individuals by vaccine type.

Twenty weeks after the second dose of vaccine, mean BNT162b2 anti-S levels were 1741 (95%CI: 1570-1931) U/ml in 18-64 year olds and 1418 (95%CI: 1317-1527) U/ml in participants over 65 years of age (Figure 2). Anti-S levels at 20 weeks post second dose of ChAdOx1 were 351 (95%CI: 320-385) U/ml in 18-64 year olds and 335 (95%CI: 308-365) U/ml in participants over 65 years of age. There was no evidence of a difference in the rates of waning by age or sex for BNT162b2 or ChAdOx1. Twenty weeks after the second dose of vaccine BNT162b2 anti-S levels were 1732 (95%CI: 1621-1850) U/ml in participants that were not clinically vulnerable, 1464 (95%CI: 1320-1623) U/ml in participants that were clinically vulnerable and 1091 (95%CI: 881-1352) U/ml in participants that were extremely clinically vulnerable. Anti-S levels at 20 weeks post second dose of ChAdOx1 were 392 (95%CI: 365-422) U/ml in participants that were not clinically vulnerable, 326 (95%CI: 293-364) U/ml in participants that were clinically vulnerable and 222 (95%CI: 177-279) U/ml in participants that were extremely clinically vulnerable. There was no evidence of a difference in rates of waning by clinical risk groups for BNT162b2 and ChAdOx1 vaccines.

8,275 individuals were included in an analysis of anti-S levels prior to breakthrough infection in order to examine anti-S as a correlate of protection against SARS-Cov-2 infection and estimate a predicted time to reach a specified anti-S level for a threshold of protection. Between 14th July 2021 and 24th October 2021, 197 individuals had a breakthrough infection. Using quartiles of SARS-CoV-2 anti-S levels, we found the risk of breakthrough infections for the lowest quartile (upper limit 413U/ml) began to diverge from the risk for higher quartiles after around 20 days of follow up (Figure 3). The risk of breakthrough infections in the 2nd lowest quartile (upper range 934 U/ml) began to diverge from that of higher quartiles after about 80 days of follow up. Informed by this descriptive analysis, we created a binary cut off of 500 U/ml for anti-S levels. Using this new categorisation of anti-S levels, we found a reduced risk of SARS-CoV-2 infection post second dose of vaccine for individuals with SARS-CoV-2 anti-S levels greater than or equal to 500 U/ml compared to those with levels under 500 U/ml (HR 0.62; 95%CIs:0.44-0.87; p=0.007; Figure 4). Using a linear mixed effect model we predicted the time to reach an anti-S threshold of 500 U/ml as 96 days for ChAdOx1 and 257 days for BNT162b2 (Figure 5).

**Figure 2.**
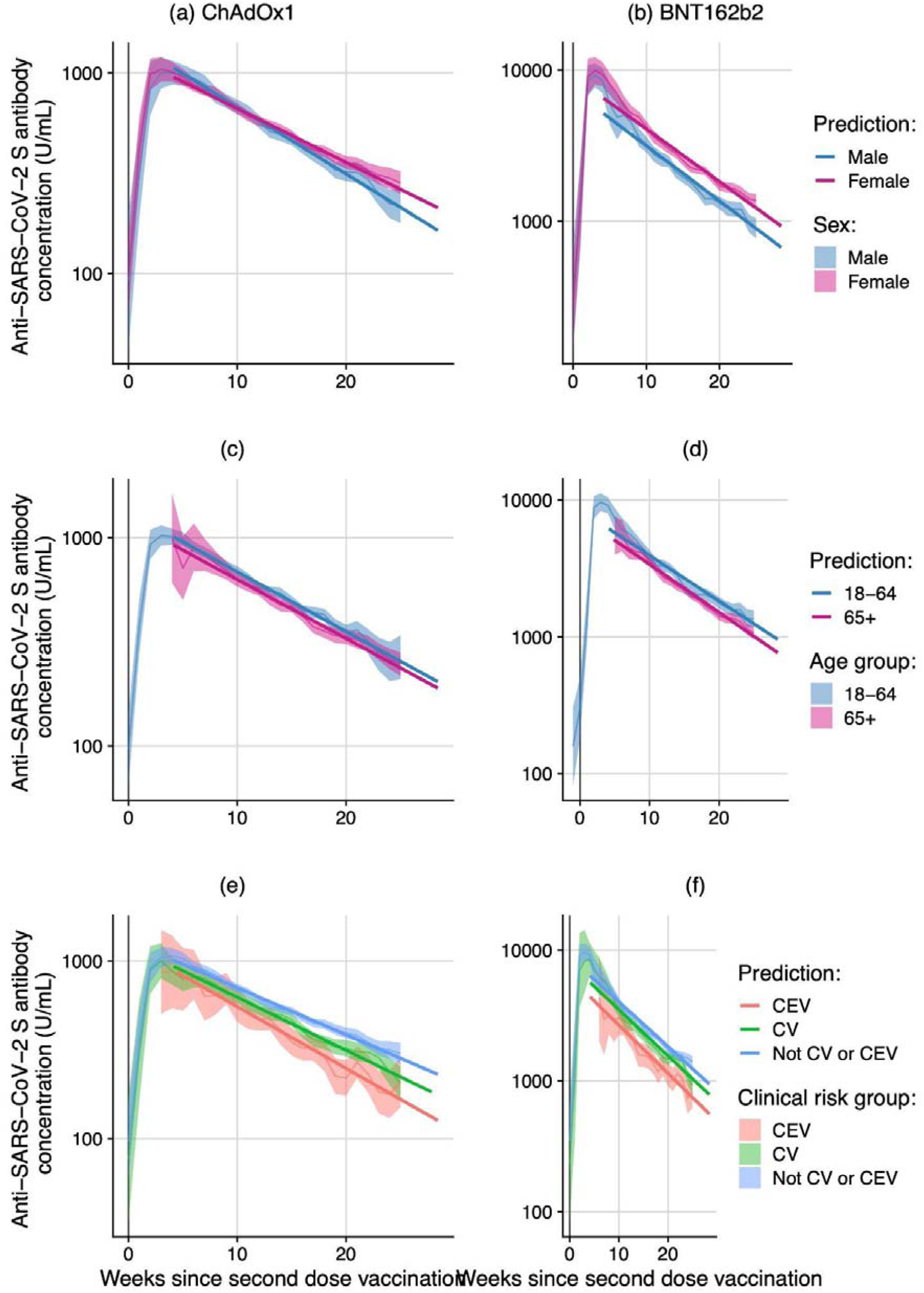
Anti-S levels (U/ml) over time since second dose of vaccination amongst N-seronegative individuals by vaccine type and sex, age and clinical risk group. ***(Note: Different y-axis scales for BNT162b2 and ChAdOx1)***

**Figure 3.**
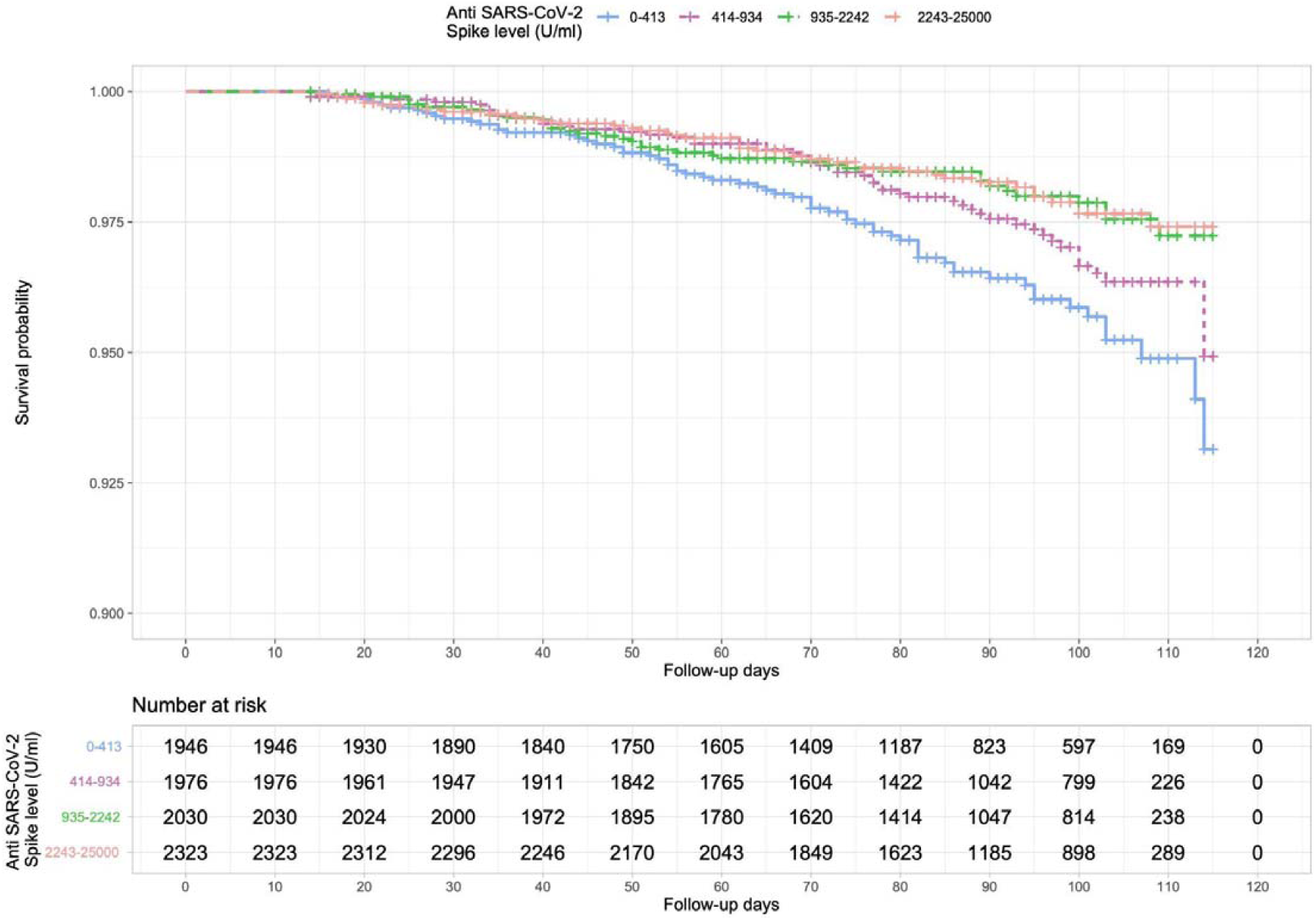
Kaplan-Meier survival curve of risk of breakthrough infection by Spike antibody level in quartiles.

**Figure 4.**
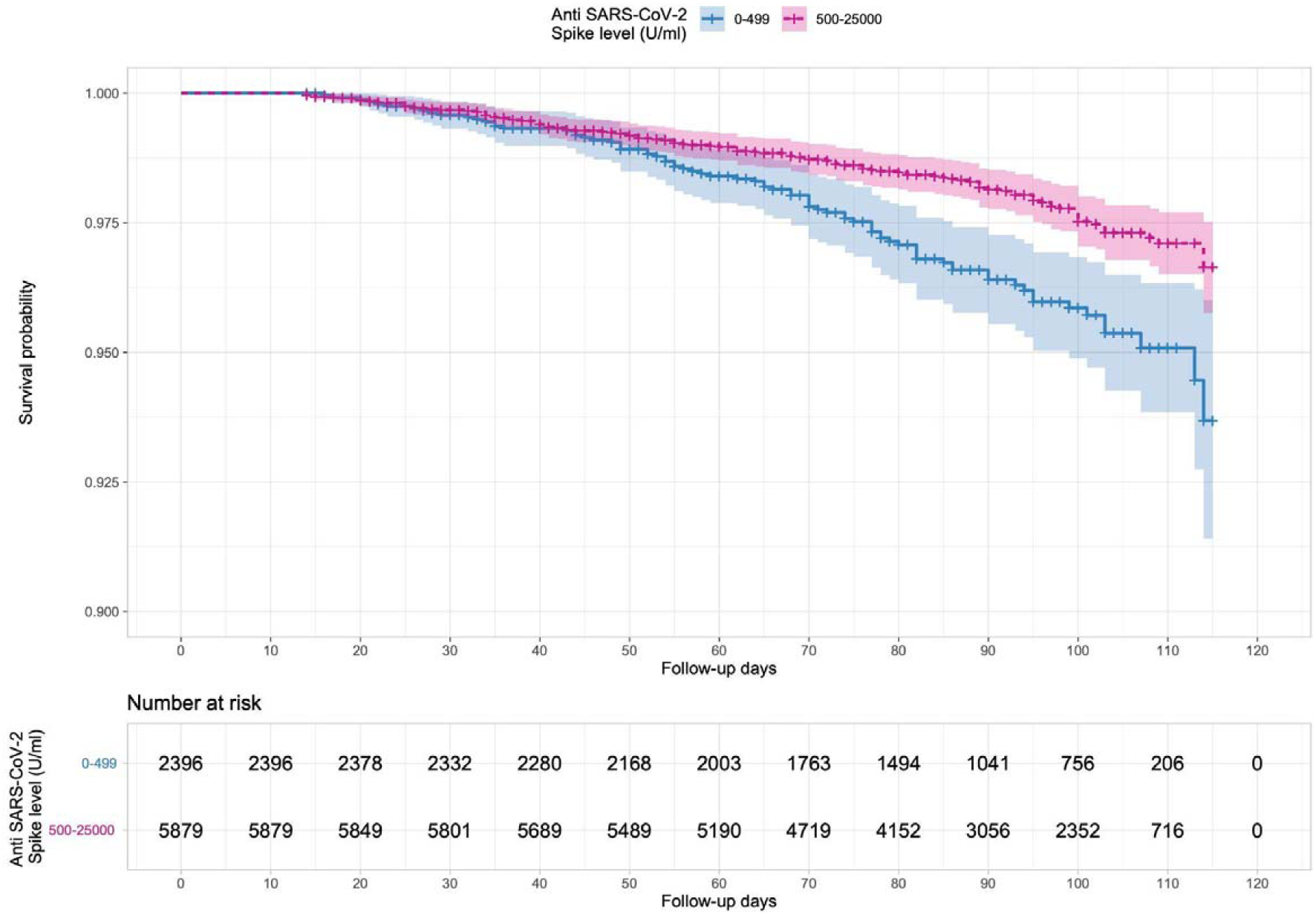
Kaplan-Meier survival curve of risk of breakthrough infection by anti-S level with binary threshold of 500.

**Figure 5.**
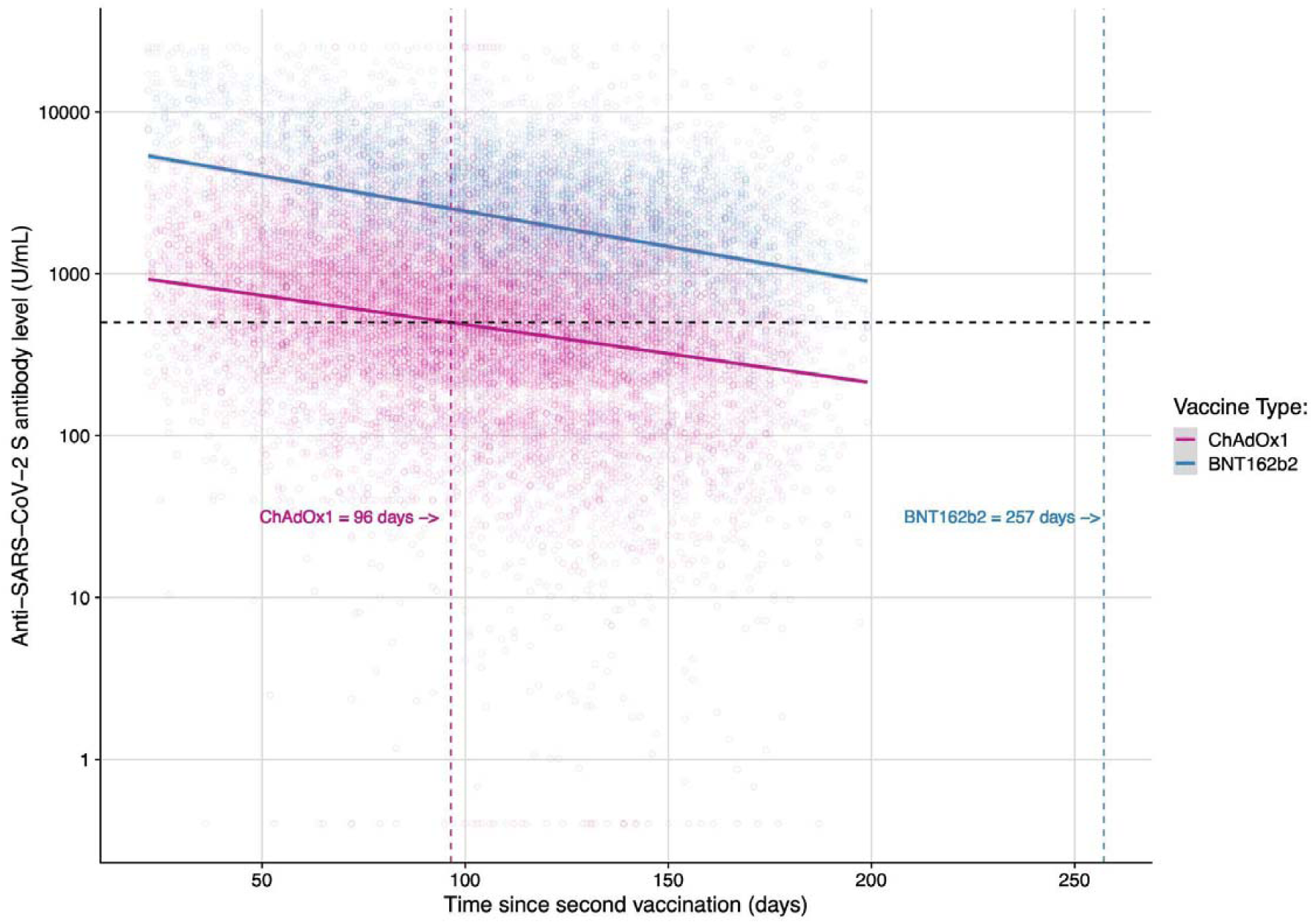
Time to anti-S threshold of 500 U/ml waning after 2nd dose of vaccine for ChAdOx1 and BNT162b2.

Participants in the test negative case-control study were drawn from 14,340 participants in the full Virus Watch cohort who reported at least one test result in the period between two weeks past the date of receiving their second vaccine dose and the date on which they received their booster dose, if applicable, or the end of study follow up. Within this set we identified 673 breakthrough infection cases occurring between 17th March 2021 and 24th October 2021 and, for each case, four matched controls (2,692) with negative test results dating between January 28 and October 24, 2021 (Table 2). We found an increased risk of a breakthrough infection for those who received the ChAdOx1 compared to those who received BNT162b2 (crude OR: 1.37, 95% CIs: 1.13-1.65, p<0.001, adjusted OR: 1.43, 95% CIs: 1.18-1.73, p<0.001).

**Table 2.**
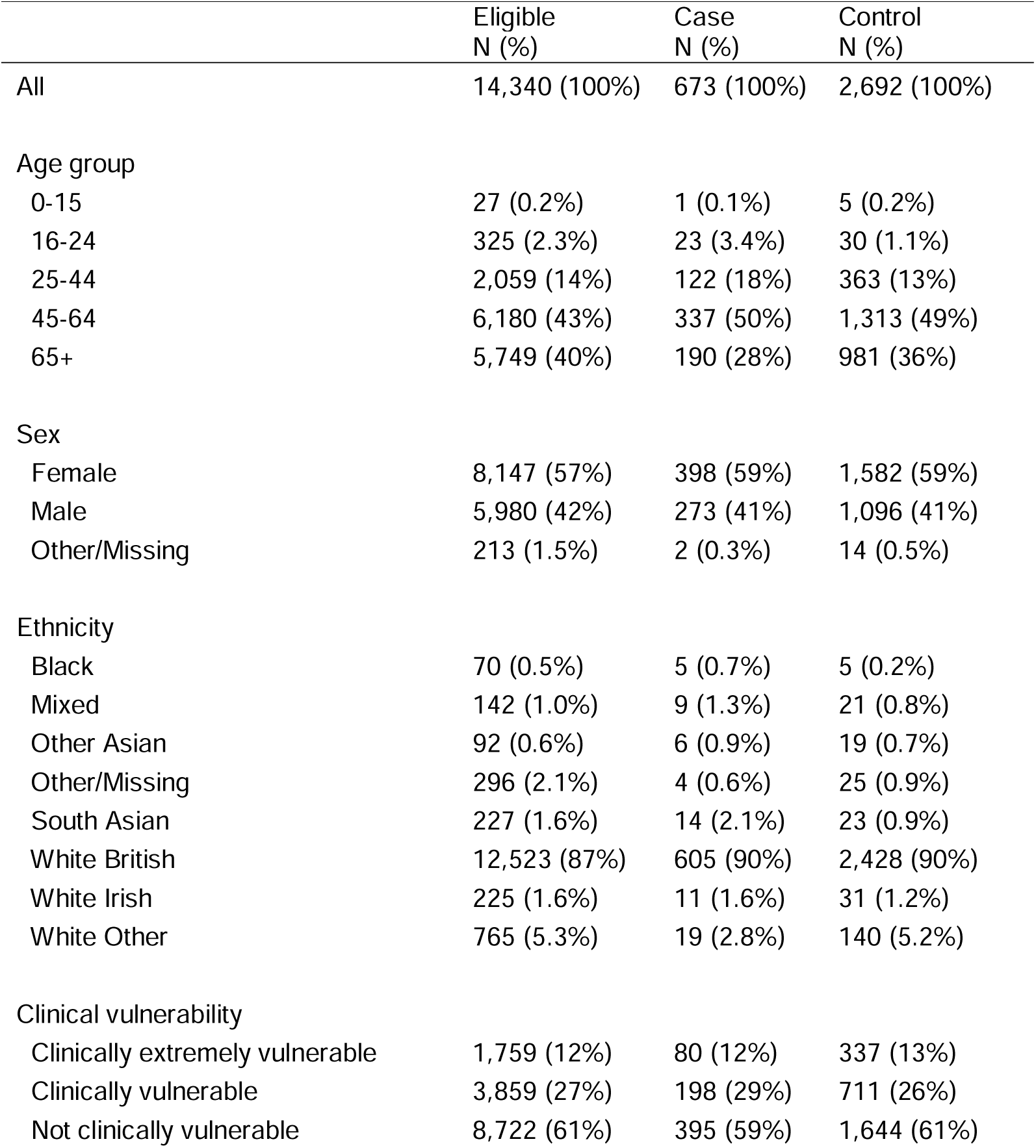
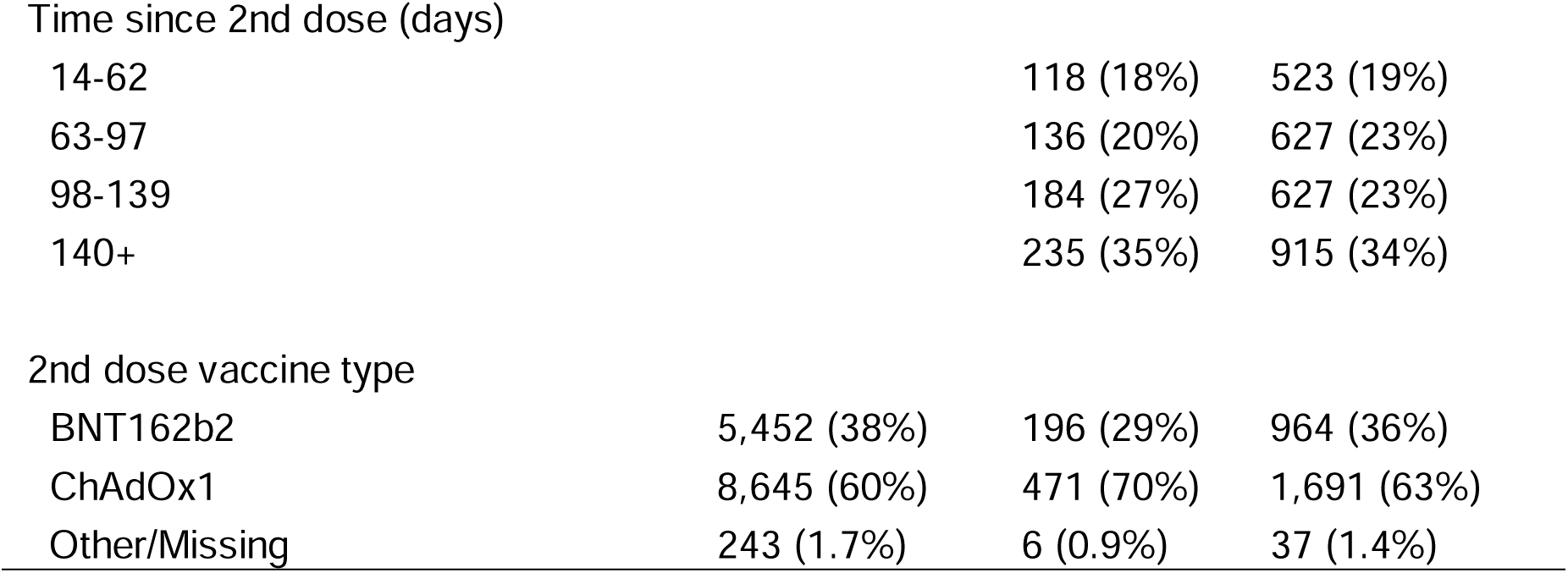
Demographic and clinical characteristics of individuals in the test negative case-control study.

## Discussion

Our study examined antibody waning in 5549 people that received a second dose of ChAdOx1 and 3205 that received a second dose BNT162b2. Anti-S levels peak following the second dose of vaccine and are nine fold higher for BNT162b2 than ChAdOx1. There is substantial waning of anti-S following both vaccines with declines following a log-linear course. We investigated 197 cases of breakthrough infection and found that those with anti-S levels of < 500 U/ml following the second dose were nearly twice as likely to have a breakthrough infection compared to those with higher levels. Mean antibody levels decline to this threshold of 500 u/ml after about 3 months for ChAdOx1 and after about 7 months for BNT162b2. These data on waning anti-S suggest those vaccinated with ChAdOx1 are likely at greater risk of breakthrough SARS-CoV-2 infection which we examined further using a test negative case control analysis from the wider Virus Watch cohort. In this analysis we found that people that had received two doses of ChAdOx1 had 1.43 increased Odds of a breakthrough infection compared to those doubly vaccinated with BNT162b2 after we controlled for time since vaccination and a range of demographic and clinical risk factors.

Strengths of our analysis include it’s community sample design from across England and Wales, with diversity in terms of age, sex and geographical location. We have repeated anti-S levels on a monthly basis from the cohort and levels were measured prior to breakthrough infection with an appropriate time window placed between SARS-Cov-2 infections and anti-S levels to ensure a clear direction of effect with our estimates. The timing of SARS-CoV-2 infections included in the study (14th July 2021 to 24th October 2021) represents a period when Delta was the main circulating strain in England and Wales, which means that our results are relevant to many countries who are currently seeing an increase in breakthrough infections due to Delta and where their populations have received ChAdOx1 or BNT162b2 vaccines. We have also been able to examine the impact of clinical risk factors on anti-S levels in our analyses and control for these in our comparative evaluation of ChAdOx1 and BNT162b2 vaccines. Since ascertainment of breakthrough infections relied on self reported tests and linked data there will have been underascertainment of breakthrough infections, and test negative designs have the advantage of being unbiased by differences in testing behaviour in different exposure groups. As the Virus Watch cohort was not randomly sampled, our measures of absolute risk of infection should be interpreted with caution when generalising to other locations, and at present we do not have data on severe outcomes e.g. hospitalisation or death. Our analysis used anti-S levels at the start of participant follow up from 1st July 2021 with breakthrough infections only included if they were 14 days post the anti-S test, therefore the threshold level associated with infection immediately prior to infection is likely to be under 500 U/ml. As further data accumulate we will be able to report on antibody levels immediately prior to breakthrough infections to better understand thresholds for protection.

Other longitudinal studies^18^ have found that antibody levels six months after the second dose of BNT162b2 vaccination decreased on average to 7% of their peak, and long term follow up studies of vaccine trial participants have found similar levels of decline and increased risk of breakthrough infection to our findings.^19^ Using data from COVID-19 vaccine trials, previous studies have estimated that neutralising antibodies to SARS-Cov-2 halve every 108 days^5^ and that a vaccine effectiveness of 90% protection against infection would decline to 70% after 6 or 7 months.^20^ Our data are consistent with these findings and provide additional information on peak anti-S levels and trajectories of waning after the second vaccination with both BNT162b2 and ChAdOx1 vaccines. A range of studies using national surveillance data linked to healthcare records from the UK, Israel and Qatar have been used to monitor BNT162b2 and ChAdOx1 vaccine effectiveness, with a consistent picture that suggests levels of protection to SARS-Cov-2 infection that wanes over time with a decline that is greater for ChAdOx1 than BNT162b2.^1–4,21^ A recent study examined the effectiveness of ChAdOx1 versus BNT162b2 COVID-19 vaccines in health and social care workers in England and found no difference in the risk of SARS-CoV-2 infection or COVID-19 disease up to 20 weeks after first dose of vaccination by vaccine type.^22^ Our data support this finding up to 20 weeks, but suggest that from this point onwards the risk of breakthrough infection for BNT162b2 is approximately half of ChAdOx1, and our anti-S levels may give an indication of the underlying correlates for this increased risk.

It is important to note that circulating anti-S levels are unlikely to be the only immunological correlate of protection against COVID-19 infection and severe disease. T-cell-mediated immunity may be particularly important^23^ and it is possible that such T-cell responses provide some continued protection against infection and severe disease as antibody responses wane. Memory B cells and T cell based immunity may also provide more stable protection,^24,25^ but remain challenging to measure at the scale needed to assess the association with protection over long time periods in prospective cohorts and prior to breakthrough infection in sufficient numbers of participants.

Overall these data show marked waning of anti-S levels over time since vaccination and provide evidence that antibody levels are an important correlate of protection against infection. The fact that anti-S levels start at a much higher level for BNT162b2 than ChAdOx1 means that breakthrough infections are likely to occur significantly earlier in those vaccinated with ChAdOx1 than BNT162b2. Our results showing an increased risk of breakthrough infections for those who were vaccinated with ChAdOx1 compared to BNT162b2 are in line with this hypothesis. This finding is consistent with national and international data showing faster waning of protection against infection and, to a lesser extent, severe disease, for ChAdOx1 than BNT162b2.

The results support the need for booster programmes prioritised to the elderly and clinically vulnerable who, because of their high risk of severe disease, were prioritised for vaccination in many countries and so have had the longest for antibody levels to decline. In the UK, those aged over 50, those who are clinically vulnerable, and healthcare workers are eligible for boosters from 6 months after their second vaccine. Our results show waning to levels associated with breakthrough infections before this 6 month period for those vaccinated with ChAdOx1 but not for BNT162b2. We also show higher risk of breakthrough infections in those vaccinated with ChAdOx1. Together, these findings suggest that boosting ChAdOx1 earlier than six months after the second dose may be advantageous, particularly in those at greatest risk of severe outcomes.

## Data Availability

We aim to share aggregate data from this project on our website and via a “Findings so far” section on our website - https://ucl-virus-watch.net/. We will also be sharing individual record level data on a research data sharing service such as the Office of National Statistics Secure Research Service. In sharing the data we will work within the principles set out in the UKRI Guidance on best practice in the management of research data. Access to use of the data whilst research is being conducted will be managed by the Chief Investigators (ACH and RWA) in accordance with the principles set out in the UKRI guidance on best practice in the management of research data. We will put analysis code on publicly available repositories to enable their reuse.

## Author contributions

Conceptualization (RWA, AY, JK, VN, SB, TB, WLEF, CG, PP, MS, IB, AMDN, AH), Data Curation (RWA, EF, VN, SB, TB, WLEF, CG, AMDN), Formal Analysis (RWA, ME, AY), Investigation (All), Methodology (All), Project Administration (JK), Resources (All), Writing – Original Draft Preparation (RWA, AY, AH), Writing – Review & Editing (All).

## Acknowledgements

We would like to acknowledge Nick Kennedy at Royal Devon and Exeter NHS Foundation Trust for the idea of plotting geometric mean data as presented in Figures 1 and 2.

