## Supplementary Appendix for "Waning of SARS-CoV-2 antibodies targeting the Spike protein in individuals post second dose of ChAdOx1 and BNT162b2 COVID-19 vaccines and risk of breakthrough infections: analysis of the Virus Watch community cohort"

**Clinically extremely vulnerable**

Individuals were categorised as extremely clinically vulnerable using criteria set out by Public Health England and the Department of Health and Social Care as part of the guidance for shielding (<https://www.gov.uk/government/publications/guidance-on-shielding-and-protecting-extremely-vulnerable-persons-from-covid-19>), which were adapted in line with clinical variables collected through the Virus Watch baseline survey, as follows:

| **Clinically extremely vulnerable (CEV) criteria as per PHE/DHSC** | **Inclusion in Virus Watch CEV definition** |
| --- | --- |
| Solid organ transplant recipients | Included |
| Cancer undergoing active chemotherapy | Included |
| Cancers undergoing radical radiotherapy | All radiotherapy included (radical radiotherapy was not ascertained) |
| Cancer of blood or bone marrow | Included |
| Immunotherapy or antibody treatments for cancer | Included |
| Targeted cancer therapies affecting the immune system | Included |
| Bone marrow or stem cell transplant in last 6 months or still taking immunosuppressive drugs | Included |
| Severe respiratory conditions including all cystic fibrosis, severe asthma and severe chronic obstructive pulmonary disease (COPD) | Included |
| Rare diseases that significantly increase the risk of infections (such as severe combined immunodeficiency (SCID), homozygous sickle cell disease) | Included |
| Immunosuppressive therapies sufficient to significantly increase risk of infection | Included |
| Problems with spleen, including splenectomy | Included |
| Down’s syndrome | Not included in CEV as not distinguished from other learning disabilities. |
| Chronic kidney disease Stage 5 or on renal dialysis | All CKD was included (stage was not ascertained) |
| Pregnancy with significant heart disease | Included |
| Others classified as clinically extremely vulnerable | Included |

**Clinically vulnerable**

Individuals were categorised as clinically vulnerable (CV) using criteria set out by the Joint Committee on Vaccination and Immunisation (<https://www.gov.uk/government/publications/priority-groups-for-coronavirus-covid-19-vaccination-advice-from-the-jcvi-30-december-2020>), excluding those who met the superseding clinically extremely vulnerable (CEV) criteria. Clinical vulnerability criteria were adapted in line with clinical variables collected through the Virus Watch baseline survey, as follows:

| **Clinically vulnerable (CV) criteria as per JCVI** | **Inclusion in Virus Watch CV definition** |
| --- | --- |
| chronic respiratory disease, including chronic obstructive pulmonary disease (COPD), cystic fibrosis and severe asthma | Included, except those that met CEV criteria |
| chronic heart disease (and vascular disease) | Included |
| chronic kidney disease | Included, except those that met CEV criteria |
| chronic liver disease | Included |
| chronic neurological disease including epilepsy | Included |
| Down’s syndrome | Included as part of broader learning disabilities |
| Severe and profound learning disability | All learning disabilities included (severity was not ascertained) |
| Diabetes | Included |
| Solid organ, bone marrow and stem cell transplant recipients | Not included (included in CEV) |
| People with specific cancers | Included, except those that met CEV criteria |
| Immunosuppression due to disease or treatment | Included, except those that met CEV criteria |
| Asplenia and splenic dysfunction | Not included (included in CEV) |
| Morbid obesity | Included |
| Severe mental illness | Included |
